# Vocabulary relearning in aphasia is supported by hippocampal memory and cortical language systems

**DOI:** 10.1101/2022.10.13.22280171

**Authors:** Katherine R. Gore, Anna M. Woollams, Matthew A. Lambon Ralph

## Abstract

Speech and language therapy can be an effective tool in improving language in post-stroke aphasia. Despite an increasing literature on the efficacy of language therapies, there is a dearth of evidence about the neurocognitive mechanisms that underpin language re-learning, including the mechanisms implicated in neurotypical learning. Neurotypical word acquisition fits within the idea of Complementary Learning Systems, whereby an episodic hippocampal system supports initial rapid and sparse learning, whilst longer-term consolidation and extraction of statistical regularities across items is underpinned by neocortical systems. Therapy may drive these neurotypical learning mechanisms, and efficacy outcome may depend on whether there is available spared tissue across these dual systems to support learning.

Here, for the first time, we utilised a reverse translation approach to explore these learning mechanisms in post-stroke aphasia, spanning a continuum of consolidation success. After three weeks of daily anomia treatment, 16 patients completed a functional magnetic resonance imaging protocol; a picture naming task which probed (i) premorbid vocabulary retained despite aphasia, (ii) newly re-learned treated items and (iii) untreated/unknown and therefore unconsolidated items. The treatment was successful, significantly improving patients’ naming accuracy and reaction time post-treatment. Consistent with the Complementary Learning Systems hypothesis, patients’ overall naming of treated items, like that of controls when learning new vocabulary, was associated with increased activation of both episodic and language regions. Patients with relatively preserved left hemisphere language regions, aligned with the control data in that hippocampal activity during naming of treated items was associated with lower accuracy and slower responses – demonstrating the shifting division of labour from hippocampally-dependent new learning towards cortical support for the efficiently-named consolidated items. In contrast, patients with greater damage to the left inferior frontal gyrus displayed the opposite pattern (greater hippocampal activity when naming treated items was associated with quicker responses), implying that their therapy-driven learning was still wholly hippocampally reliant.

**Open access:** For the purpose of open access, the UKRI-funded authors have applied a Creative Commons Attribution (CC-BY) licence to any Author Accepted Manuscript version arising from this submission.

## Introduction

Around one third of people develop aphasia after cerebral stroke.^1^ The most common symptom of post-stroke aphasia is word-finding difficulties (anomia) which often persist into the chronic phase of aphasia recovery. Anomia severely reduces patients’ communication and, therefore, quality of life.^2^ Thus, successful treatment of anomia is paramount. Although there is extensive evidence that speech and language therapy (SLT) can be effective^3–5^, the underlying neural correlates of therapy effectiveness and its variability remain unclear. There have been few, if any, studies that compare SLT-induced changes in neural activation with those associated with neurotypical mechanisms of vocabulary learning. In this reverse translation study, therefore, we utilised post-intervention fMRI to reveal the neural correlates of successful naming therapy, explored how these relate to the neural correlates of vocabulary learning in neurotypical participants, and investigated how lesions to core left hemisphere language regions change the neural division of labour that supports vocabulary relearning.

Previous studies have explored treatment-induced changes in brain activity^6–8^, and considered how SLT might induce neuroplasticity^9^, the proposed explanations of which have tended to mirror the hypotheses and debates about spontaneous recovery of language function in aphasia. Thus, some studies have proposed that recovery is dependent on the reconstitution of language systems in undamaged tissue around the lesion, resulting in left perilesional activity^8,10–14^. Other studies have reported a shift of language function to the contralateral hemisphere, with increased activity in right language homologues.^15–17^ Accordingly, there is extensive debate about which patterns of reorganisation are advantageous to language outcome in stroke: the contribution of right hemisphere language homologues to speech production has been considered both beneficial^15,16,18^ or detrimental^19,20^ to recovery. Recent neurocomputational models suggest that these apparently contradictory patterns actually reflect the process of recovery after different levels of damage within an asymmetric yet inherently bilateral language production system.^21^ There is also some evidence that, beyond language-specific brain regions, there can be treatment-induced functional changes in domain-general brain networks^22^, including networks underpinning short-term memory.^23,24^

An important next step in the field would be to secure a neurocognitive theory of SLT-induced language improvements, ideally underwritten by formal computational models. A first stride in this direction might be derived from the rich literature on learning and memory consolidation in healthy individuals. Explorations of the neural processes associated with word training in healthy participants align with the Complementary Learning Systems theory (CLS).^25–28^ The CLS model proposes that sparse and highly plastic representations within the medial temporal lobes (MTL) and other parts of the episodic network support initial rapid knowledge acquisition. Slower learning across distributed representations within the neocortex allows for extraction of statistical structures and generalisation across episodes. Over time, knowledge consolidation is driven by a gradual MTL-to-neocortical shift in the division of labour across these complementary systems.^25^ Previous fMRI investigations have shown that initial exposure to novel, abstract vocabulary items engages the hippocampus which gradually reduces over repeated presentations.^29–32^ A more recent study in healthy participants, which adopted the typical multi-session cueing-based learning/therapy approach used for aphasic patients in SLT, investigated the neural division of labour for consolidated learning of previously-unknown native vocabulary items.^28^ Consistent with the CLS theory, the newly-consolidated vocabulary was reliant upon both episodic regions (including MTL) and neocortical language regions; by contrast, pre-existing vocabulary only engaged these neocortical language regions. In line with the shifting MTL-to-neocortical division of labour, participants’ language efficiency with these items was positively correlated with activation in the neocortical regions and negatively related to the hippocampal activation.

In this reverse translational study, we investigated the underlying neural correlates of successful speech and language therapy. By following the design of the recent exploration of native vocabulary learning in healthy participants (see above), we addressed two central questions: (1) does word re-learning in post-stroke aphasia fit within the CLS framework that supports word learning in healthy older adults?; and (2) does a certain level of critical damage to the core IFG speech production area cause deviations away from this typical learning framework? To explore these questions, we gave patients three weeks of speech and language therapy on items they could not consistently name in prior naming assessments. Directly after treatment, patients completed a picture naming task in the fMRI scanner in which we compared the activation associated with naming of treated items, untreated items they could name prior to scanning, and untreated/unknown items.

By mirroring the procedure of the healthy control study^28^, we were able to compare the patients’ fMRI results for name relearning against the combined language and episodic networks that are observed for vocabulary learning in healthy participants. In advance of the study, we anticipated three possible outcomes: (i) if enough of the patient’s typical language systems were intact, then the naming of these relearnt items should mirror the healthy control pattern; (ii) if representations of the item’s name exist but were weakened following stroke, re-learning could strengthen the representations and processing in the typical (retained) language system. This ‘representational repair’ process may be supported by ‘new learning’ via the episodic system, similar to when controls learn new vocabulary^28^; (iii) if language areas were more compromised, then the remaining representations may be more severely damaged and thus (a) compensatory language-related systems may be engaged, or (b) if the neocortical representations are critically compromised then episodic systems would be dominant, somewhat akin to the pattern observed in healthy individuals during the initial phase of novel vocabulary learning^29–32^ where there are yet to be any cortical representations.

## Materials and methods

### Participants

Sixteen participants were recruited from the North West of England (seven female, age range = 39-73, mean (*M)* = 57.5) with acquired language production impairment following a single left hemispheric stroke (either ischaemic or haemorrhagic) at least one year prior to the study (see Figure 1 for lesion overlap and Table 1 for demographic and selected neuropsychological data). Participant numbers were determined based on access. Inclusion criteria were: (i) anomia as determined by the Boston Naming Test (BNT; cut off 90%)^33^; (ii) good spoken comprehension determined by the Comprehensive Aphasia Test (CAT)^34^; (iii) good repetition of single words and non-words determined by the Psycholinguistic Assessments of Language Processing in Aphasia (PALPA)^35^ and (iv) absence of apraxia of speech. Participant’s aphasia was classified using the Boston Diagnostic Aphasia Examination (BDAE).^36^ All were native English speakers, with normal or normal-to-corrected vision and hearing, with no other significant neurological conditions or contraindications to MRI scanning. Participants gave informed consent prior to participation according to the Declaration of Helsinki, with approved ethics by the local Health Research Authority ethics committee. Structural MRI data from an age and education matched healthy control group (*n* = 20, 12 females, age range = 46-77, *M* = 63.90) were used for lesion identification.^37^

**Figure 1.** Lesion overlap map of the 16 participants on axial slices. Colour bar indicates the number of participants included in the overlap. Numbering above slices indicates *z* MNI coordinate. In neurological convention, left is left.

**Table 1.**
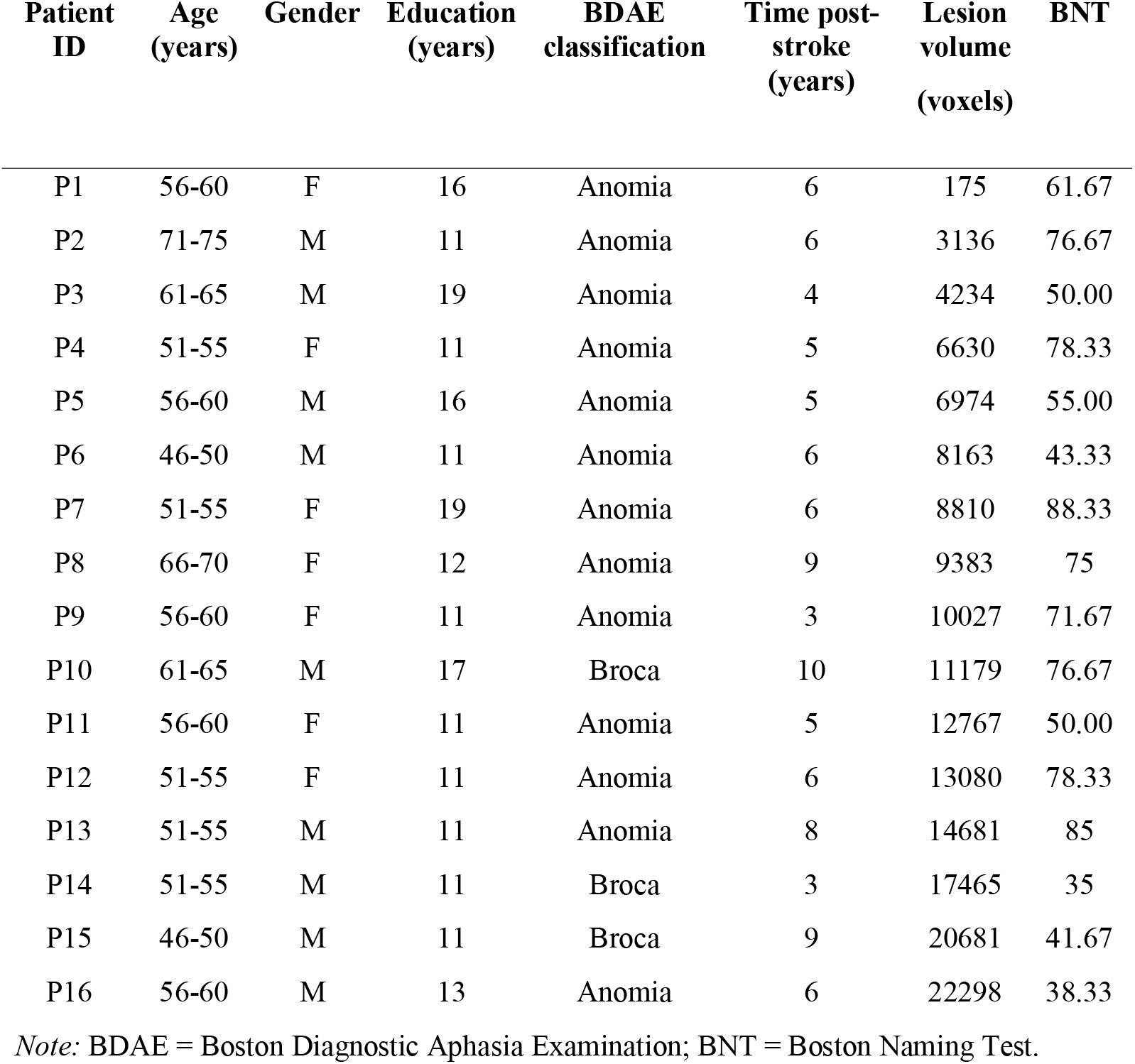
Patient’s demographic information ordered by lesion volume.

### Background neuropsychological assessments

All participants were tested on an extensive neuropsychological/aphasiological battery assessing language and cognitive abilities.^38^ In addition to the BDAE^36^, this battery consisted of (i) naming tests, including the Boston Naming Test (BNT^33^) and the 64-item Cambridge naming test; (ii) the spoken sentence comprehension task from the CAT^34^; (iii) semantic tasks from the 64-item Cambridge Semantic Battery^39^ and the 96-trial synonym judgement test^40^; (iv) speech production tests from the PALPA^35^; (v) cognitive tests including the forward and backward digit span,^41^ Raven’s Coloured Progressive Matrices^42^ and the Brixton Spatial Rule Anticipation Task^43^. Patient’s demographic details are displayed in Table 1, alongside results of the BNT to demonstrate that there was a word-finding deficit. An overlap map of participants’ left hemispheric lesions is shown in Fig. 3C.

### Pre-treatment naming assessments and stimuli

The design mirrored that used in Gore et al.^28^ with healthy controls. Patients were assessed on all stimuli items three times prior to training. There were three sets of items: already known, to-be-treated and untreated. 200 items were taken from the International Picture Naming Project^44^ and matched with discernible, coloured images with a white background to build the already known and to-be-treated item sets. Items were selected with high word frequency, short reaction times (<1000ms) and high accuracy (85-100%) in the normative IPNP data. Items that were correctly named by a patient on all occasions were added to that individual’s already known set. Items that could not be named on one or more occasions were randomly divided between the to-be-treated or untreated sets. Final sets comprised 50 items each and were matched across sets for their psycholinguistic properties of word frequency, number of syllables and word length. Participants who could name over 80% of all items before training were provided with a more challenging set of to-be-treated items (3 participants in total). This set contained unfamiliar, rare items with low word frequency names, drawn from the British National Corpus.^45^ Images from the already known, treated and untreated sets were phase scrambled for use as stimuli in the baseline task.

### Anomia treatment

Patients received a laptop to practice naming the to-be-treated items for 45 minutes a day, four days a week, for three weeks. Patients completed, on average, a total of 7.3 hours of naming treatment. During the first two weeks, participants received cue-based training. In the third week, participants received speeded training.

During cue training, the number of items was incrementally increased, beginning with 10 items. When 70% accuracy was reached on these 10 items, another 10 items were added incrementally up to the total 50 items. Participants first viewed each item picture and item name in orthographic and audio form. The patients repeated the name out loud. The cue training was similar to standard error-reducing speech and language therapy ^46,47^, but with a choice of cue to provide a self-determined approach. An item picture was shown, and patients had an option to choose cues or to proceed with naming. For example, for a picture of a butterfly, the cue options were (i) written semantic description “A flying insect with large colourful wings”; (ii) an initial phonemic cue ‘bu’; (iii) initial and second phonemic cue ‘butter’ and (iv) the whole word cue ‘butterfly’. All cues were delivered orthographically and audibly by the laptop. After each naming attempt, the whole correct word was given. Patients then answered a semantic question “Is it living?”.

The third week of training adopted the repeated increasingly-speeded presentation (RISP^48^) method. To emphasise speed as well as naming accuracy, patients were instructed that they needed to name the item before the computer did. When participants reached a speeded naming success rate of 70%, the timing was incrementally decreased from 3s to 1.4s, to 1s. When participants beat the 1s target for 70% of items, the set size was increased by 10 items and the time-to-beat returned to 3s.

### Procedure

Patients were asked to name the 200 IPNP pictures without cues before and after the 3-week anomia treatment. Within two days of finishing treatment, patients performed a picture naming task in the scanner. Patients also performed a semantic task, which is the focus of another study. The order of pictures and the order of task was counterbalanced within and across patients.

### Neuroimaging acquisition

High-resolution structural T1-weighted MRI scans were acquired on a 3T Philips Achieva scanner using an eight-element SENSE head coil. The parameters were as follows: repetition time = 9.0ms, echo time = 3.93ms, acquired voxel size = 1.0 × 1.0 × 1.0 mm squared, matrix size = 256 × 256, slice thickness = 1mm, flip angle = 8 degrees, FOV = 256mm, inversion time = 1150ms, with 150 contiguous slices and a SENSE acceleration factor of 2.5.

A triple gradient echo EPI sequence was used for functional imaging to improve signal-to-noise ratio in the anterior temporal lobes (ATL).^49,50^ In single echo imaging, there is signal dropout and distortion in the ATL.^51–53^ The functional scans were also acquired with a 45-degree tilt from AC-PC to reduce ghosting artefacts in the temporal lobes. The functional sequences consisted of 31 slices covering the whole brain, TR = 2.5s, TE = 12, 30 and 48ms, flip angle = 85 degrees, resolution matrix = 80 × 80, FOV = 240 × 240mm and voxel size = 3.75 × 3.75 × 4mm.

Stimuli were presented during scanning using E-Prime 2.0, with block order pseudo-randomised and optimised for statistic power using OptSeq (http://surfer.nmr.mgh.harvard.edu/optseq/). Spoken in-scanner responses were recorded using a fibre optic, noise-cancelling microphone for fMRI (FOMRI; Optoacoustics). Patients practised speaking with as little movement as possible prior to scanning to reduce motion artefacts.

There were two tasks during imaging acquisition, one of which is the focus of a separate study. For this study, a blocked-design picture naming task with four conditions was used; already known, treated, untreated and baseline. In the known condition, patients named pictures they could consistently name prior to treatment (e.g., pencil). In the treated condition, patients named newly treated items (e.g., penguin). If patients could not remember an item name, or the item was novel or phase scrambled they responded: “Don’t know”.

Each trial lasted 3700ms. A fixation cross was presented for 700ms, followed by the image to be named for 3000ms. Each 11.1s block consisted of three trials. Eight jittered length rest blocks were included per run, with an average length of 11.1s. There were two 7 minute and 4 second long blocks per specific task.

### Neuroimaging preprocessing and analysis

T1-weighted structural images were pre-processed in FSL, version 6.0.0,^54^ using the fsl_anat pipeline (http://fsl.fmrib.ox.ac.uk/fsl/fslwiki/fsl_anat), excluding FAST tissue segmentation. A supervised lesion segmentation algorithm (LINDA) was used for lesion identification.^55^ T1-weighted images were normalised to Montreal Neuroimage Institute (MNI) space and LINDA transforms were applied. The functional data were despiked and slice time corrected per echo in the AFNI neuroimaging suite (v19.2.10; 3dDespike; 3dTshift).^56,57^ Motion correction parameters were calculated from the first slice of the first echo, then this transformation was applied to all three echoes. A subject-specific, LINDA-generated T1 mask was transformed to functional space and applied to each echo. Multi-echo data were optimally combined using the T2* combination method^58^ in tedana.^49,59^ Optimally combined data were normalised to MNI space using LINDA transforms. Smoothing was performed on this data using FSL, using 8mm full-width half-maximum equivalent sigma. Statistical whole brain analyses were performed non-parametrically using SnPM13 (http://warwick.ac.uk/snpm). Implicit masking defaults were set to ‘-Inf’ to remove the arbitrary analysis threshold and avoid lesion voxels being masked out for all participants. Region of interest analyses were performed using SPM12^60^ and MarsBaR^61^. Two-step multiple regression and simple slope analyses were performed in R version 4.0.5^62^ using the emmeans package.

Regions of interest were defined based upon the previous literature. Five *a priori* regions of interest were defined to include key semantic and episodic network areas. The left inferior frontal gyrus (IFG; MNI: -46 28 10), a key area of damage in post-stroke aphasia is considered critical in speech production.^63–65^ The right IFG homologue (MNI: 46 28 10) was included to investigate right-sided homologue compensation theories.^66–68^ A left ventral anterior temporal lobe (vATL; MNI: -36 -15 -30) was taken from a key semantic cognition reference.^69^ For the episodic memory network, a left hippocampal ROI (MNI: -28 -14 -15) was defined based on previous literature of initial hippocampal activation during word learning.^29,30^ In addition, an ROI in the left inferior parietal lobe was included (IPL; MNI: -47-64 34) due to consistent activation of this region in episodic processing.^70,71^ Pearson’s *r* correlations were used to explore associations between ROI BOLD activity and behavioural performance measures (accuracy and RT). Critical tests of significant differences between correlations were adjusted for multiple comparisons using the Benjamini-Hochberg procedure, FDR = 0.05.^72^

To explore the interaction between critical language region damage and performance/functional activity, a further left IFG ROI was created. This ROI included left IFG pars opercularis and left IFG pars triangularis, as designated by the Automated Anatomical Labelling atlas.^73^

### Behavioural performance analysis

Individual behavioural performance was probed for differences between the three pre-training test scores using a McNemar’s test. Pre-treatment scores were averaged per participant as baseline performance was stable across the group and individuals. Repeated-measures *t*-tests were performed for each stimuli type, between the time points of pre- and post-treatment for accuracy and RT. Two repeated-measures analyses of variance (ANOVA) were conducted, one for accuracy performance and one for RT. These ANOVA analyses included within-subject main factors of Time (pre-treatment, post-treatment) and Treatment (treated, untreated and already known). Two-tailed *p* values and an alpha level of 0.05 were applied for all statistical tests. Due to the global pandemic, maintenance data was not available.

## Results

### Behavioural performance

There was no significant difference in the proportion of accurate responses between the three baseline pre-tests across the group (*p* = .88). Pre-treatment performance showed no significant change across time points. Thus, baseline pre-tests were averaged to provide a single representative sum of pre-treatment baseline performance. There was no significant difference in accuracy (*t* = 1.67, *p* = .12) or RT (*t* = -1.76, *p* = .10) for untreated items between time points (Fig. 2A). The patients demonstrated a substantial and significant improvement in naming accuracy (*t* = 5.18, *p* < .0001) and improved naming speeds (*t* = - 7.63, *p* < .0001) when naming treated items after treatment (Fig. 2B). The patients also showed a small yet significant increase in naming accuracy for the known items (*t* = 3.31, *p* = .004), but the small change in naming speed failed to reach significance (*t* = -2.08, *p* = .06; Fig. 2C).

**Figure 2.**
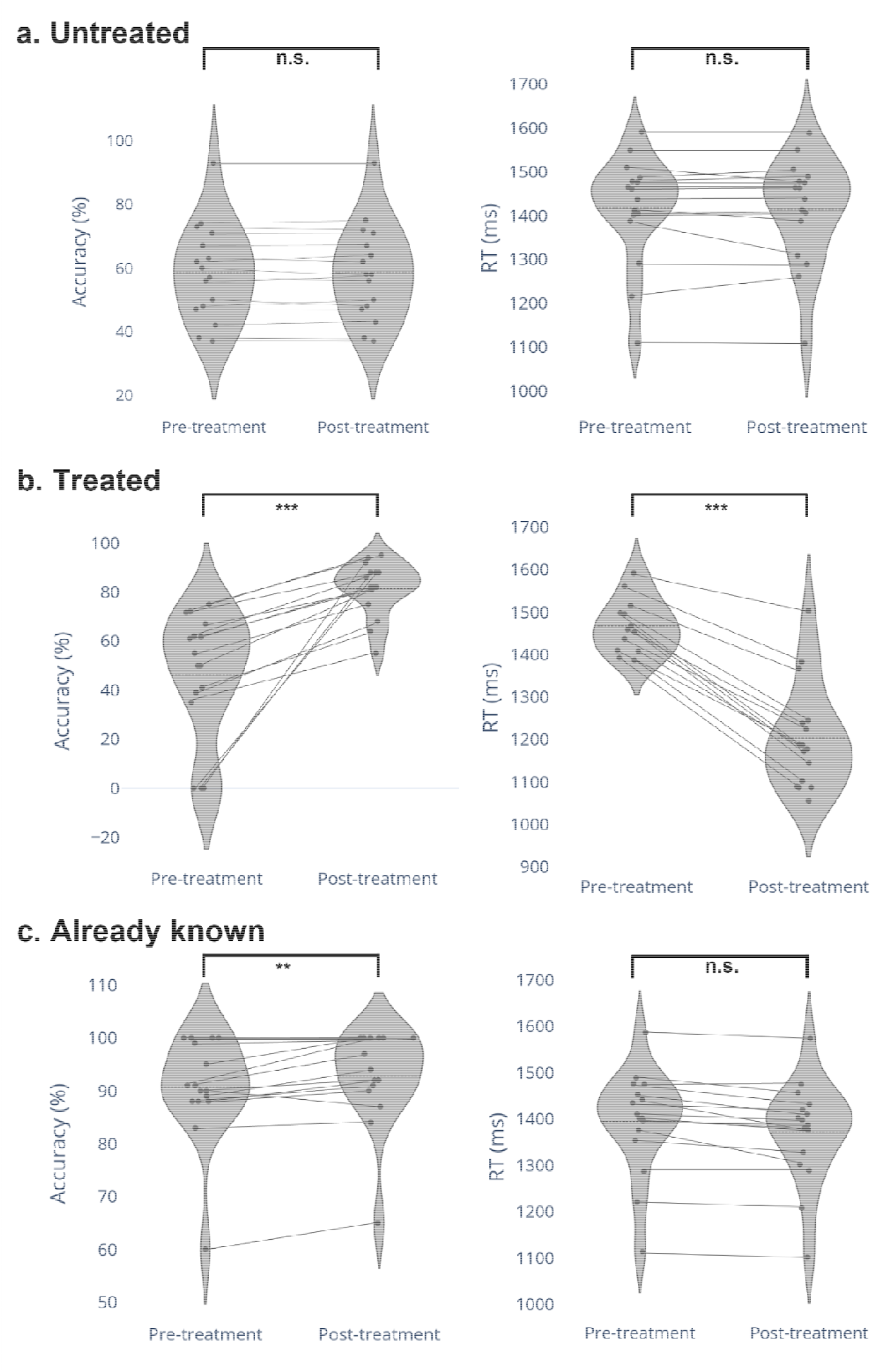
Accuracy and RT of naming a) untreated items, b) treated items and c) already known items. Dotted line represents the mean. Solid lines between points represent individual patient differences in time points. Group differences significant at p < .0001 and p < .005 are denoted with ‘***’ and ‘**’ respectively.

### Accuracy

A repeated-measures 2 × 3 ANOVA revealed a significant main effect of Time (*F* (1,15) = 90.35, *p* < .001), with greater accuracy post-treatment. There was also a significant effect of Treatment (*F* (2,30) = 118.31, *p* < .0001) with greater accuracy when naming newly treated than untreated or known items. There was a significant Time x Treatment interaction (*F* (2,30) = 35.48 *p* < .0001). Naming of treated items was more accurate post-treatment (81%) than pre-treatment (46%). This unstandardised effect size (35%) was significantly greater than the mean difference (1%) between naming of untreated items pre-treatment (58%) and post-treatment (59%; *p* < .0001). The unstandardised effect size was also significantly greater than the mean difference (2%) between naming of already known items pre-treatment (90%) and post-treatment (92%; *p* < .0001).

### RT

There were also significant main effects of both Time (quicker RTs post-treatment; *F* (1,15) = 24.084, *p* < .001) and Treatment (faster RTs when naming newly treated than untreated or known items; *F* (2,30) = 27.307, *p* < .0001). There was also a significant Time x Treatment interaction for RT (*F* (2, 30) = 17.507, *p* < .001). Naming of newly treated items was faster (1203ms) than pre-treatment naming (1467ms). This unstandardised effect size (264ms) was significantly greater than the mean difference (4ms) between untreated items pre-treatment (1417ms) and post-treatment (1413ms; *p* = .002). Additionally, the unstandardised effect size was also significantly greater than the mean difference (23ms) between already known items pre-treatment (1394ms) and post-treatment (1371ms; *p* = .002).

### Whole brain fMRI contrasts

The results of whole brain analyses are reported in Table 2. Multiple contrasts were performed; treated > already known, treated > untreated, already known > untreated and the inverse of these contrasts. There were significant clusters for treated > untreated and already known > untreated (Fig. 3). In the treated > untreated contrast, there was a significant cluster of activation over the right superior temporal gyrus (STG), right rolandic operculum and the right postcentral gyrus. There was also a further cluster in the dorsal occipital cortex spanning bilateral lingual gyri and the cerebellum. In the already known > untreated contrast, there was a similar pattern of activation. The right-sided cluster spanned the right anterior parietal and superior temporal cortex areas. The posterior cluster centred on the bilateral lingual gyri, including posterior left parahippocampal cortex.

**Table 2.** Significant clusters of activity during naming of treated, already known and untreated items.

| Contrast | Region of activation | Peak region | Cluster size | Peak MNI |  |  | Pseudo *t* |
| --- | --- | --- | --- | --- | --- | --- | --- |
|  |  |  |  | x | y | z |  |
| Treated > untreated | R STG, postcentral gyrus | R STG | 3947 | 65 | -14 | 11 | 5.26 |
|  |  | R postcentral |  | 54 | -6 | 13 | 5.05 |
|  |  | R STG |  | 64 | -6 | 4 | 3.86 |
|  | Occipital cortex, cerebellum | R cerebellum | 24708 | 17 | -59 | -23 | 5.15 |
|  |  | L calcarine |  | 2 | -86 | -5 | 4.84 |
|  |  | L cerebellum |  | -13 | -62 | -13 | 4.33 |
| Known > untreated | R SMG, R STG, R Heschl gyrus | R SMG | 2855 | 61 | -35 | 29 | 4.84 |
|  |  | R STG |  | 65 | -16 | 12 | 4.35 |
|  |  | R STG |  | 64 | -7 | 7 | 3.76 |
|  | Bilateral lingual gyrus, left MTL | L fusiform | 9039 | -31 | -39 | -11 | 4.71 |
|  |  | R lingual |  | 5 | -79 | -7 | 4.65 |
|  |  | L calcarine |  | 2 | -85 | 0 | 4.39 |

*Note:* Clusters significant at $p < .001$ voxel height and $p < .05$ FWE cluster correction. Up to 3 strongest peaks listed per cluster. Pseudo-*t* statistics were computed with smoothed variance in SnPM. R, right; STG, superior temporal gyrus; SMG, supramarginal gyrus; MTL, medial temporal lobe.

**Figure 3.**
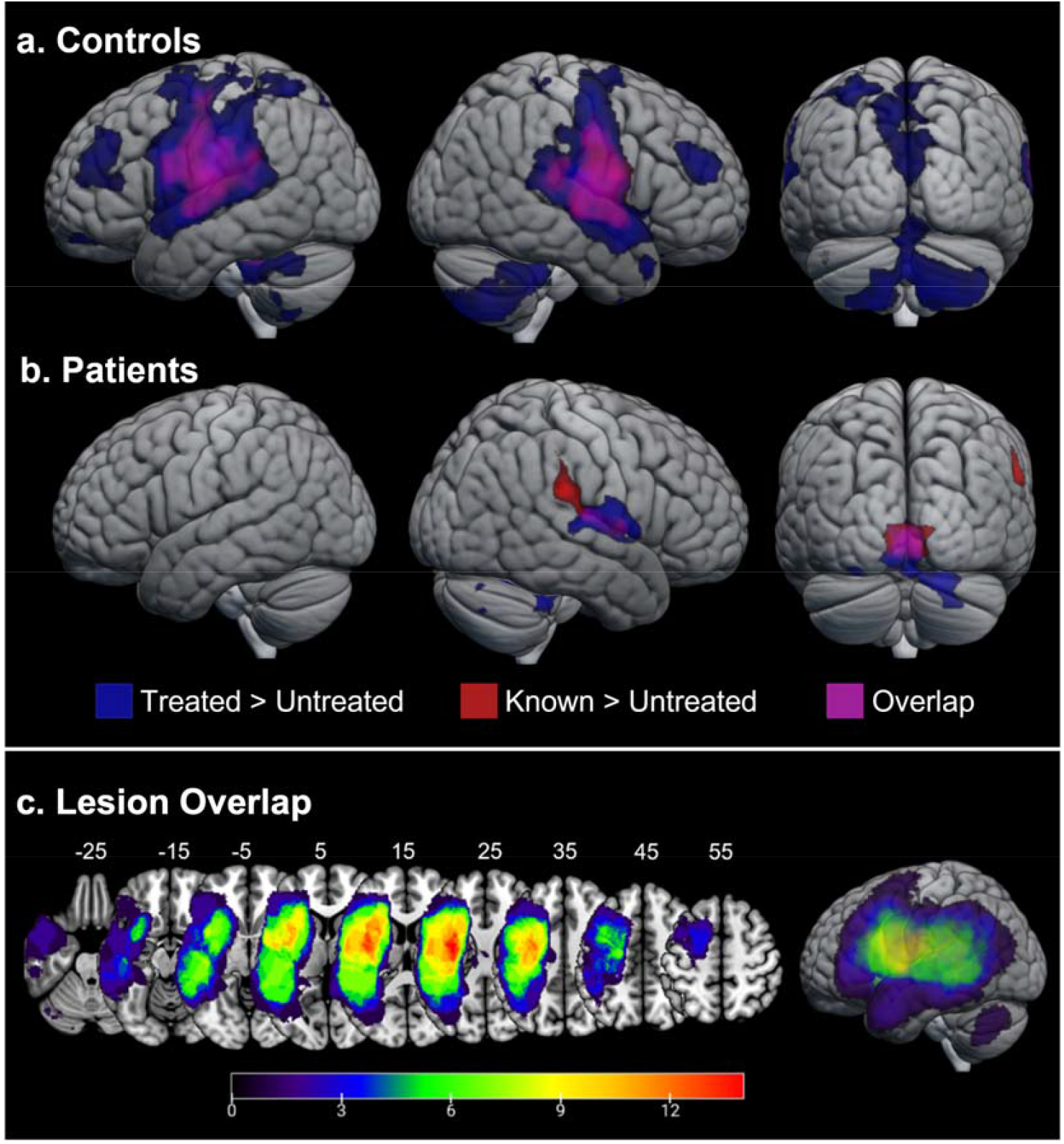
Whole brain clusters of increased activation during picture naming for (A) controls and (B) patients. Patient lesion overlap map on axial slices (C). Picture naming of treated > untreated (blue) and known > untreated (red) contrasts, with overlap (purple). Image thresholded at voxel level *p* < .001, cluster corrected FWE *p* < .05. R; right, P; posterior. (C) Colour bar indicates the number of participants included in the overlap. Numbering above slices indicates z MNI coordinate. In neurological convention, left is left.

### Region of interest analyses

To explore the core hypothesis that neural processes underlying word re-learning in aphasia would be similar to those that support word learning in healthy controls (e.g., CLS theory) five *a priori* regions of interest (ROIs) were defined (Fig. 4A). These included language-semantic network areas (left IFG, right IFG and left ATL) and episodic network areas (left hippocampus and left IPL). Percentage damage to 8mm spherical ROIs is listed in Supplementary Table 1. An initial exploratory analysis was performed correlating BOLD activity of these ROIs with naming reaction times (RT) and accuracy. There were no significant correlations between behaviour and ROI activity in the already known > untreated contrast.

**Figure 4.**
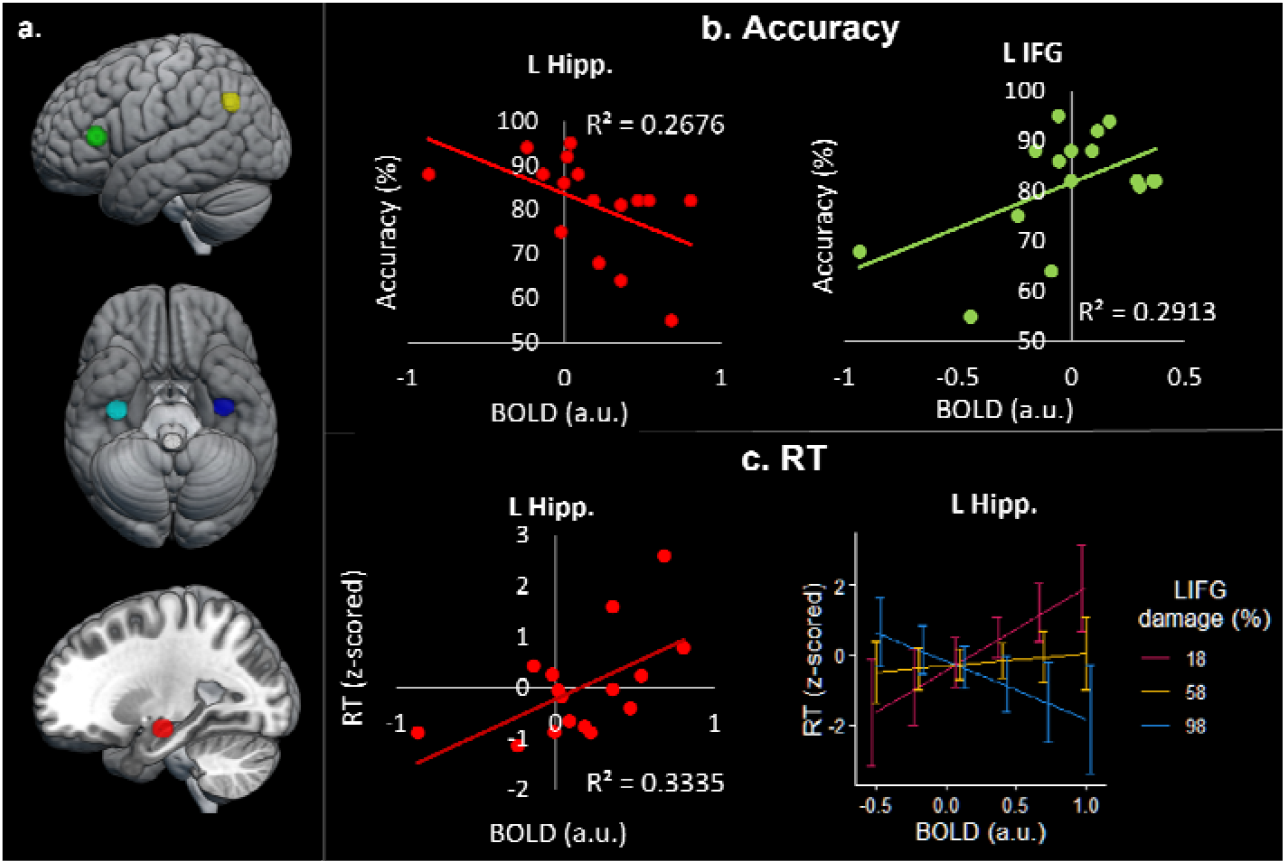
Correlations between regional activation and behavioural performance. (**A**) Regions of interest: green = left inferior frontal gyrus (IFG); yellow = left inferior parietal lobe; cyan = left anterior temporal lobe (ATL); blue = right ATL; red = left hippocampus (Hipp.). (**B**) Correlations between treated > untreated BOLD activity and accuracy. (**C**) Left: correlation between treated > untreated left hippocampal BOLD activity and RT. Right: Predicted simple slopes of the association between treated > untreated left hippocampal BOLD activity and RTs at three levels of left IFG damage.

In Gore et al.^28^ the underlying neural correlates of word learning fitted within the CLS model. To investigate whether speech and language therapy also drives these neurotypical learning mechanisms, we explored whether treated > untreated behavioural-activation correlations differed significantly from already known > untreated items. In healthy participants, left hippocampal activity was associated with worse performance, whereas left IFG and left ATL activity was associated with better performance.^28^ For the patients in this study, there was a strong negative correlation between accuracy and left hippocampal activity in the treated > untreated condition (*r* = -.517, *p* = .039, Fig. 4B), which was significantly different to the weak *positive* correlation of accuracy and left hippocampal activity in the already known > untreated condition (*r* = .363, *p* = .166) using Fisher’s r-to-z transformation (*z* = -2.43, *p* = .008, adjusted *p* = .024). There was a positive correlation between naming accuracy and left IFG activity in the treated > untreated condition (*r* = .540, *p* = .043, Fig. 4B), which was significantly different to the weak negative correlation of naming accuracy and left IFG activity in the already known > untreated condition (*r* = -.025, *p* = .929) using *r*-to-*z* transformation (*z* = 2.19, *p* = .014, adjusted *p* = .021). There were no further significant correlations between accuracy and ROI BOLD activity, including in the right IFG *r* = .041, *p* = .881.

Associations between region of interest BOLD activity and naming RTs were also explored. There was a strong positive correlation between longer RTs and left hippocampal activity in the treated > untreated condition (*r* = .576, *p* = .020. Fig. 4C), which was significantly different to the weak positive correlation in the already known > untreated condition (*r* = - 0.011, *p* = .971; *z* = -1.65, *p* = 0.04, adjusted *p* = .04). There were no further significant correlations between RT and ROI BOLD activity, including in the right IFG (*r* = .229, *p* = .260).

To examine the second study question, hierarchical multiple regression was used to investigate whether neural changes may deviate from those found in normal word learning paradigms dependent on levels of damage to critical regions. Given its importance in language processing and speech production, we focussed on the level of damage to the left IFG. All covariates were mean centred, and the interaction term was generated. In the first step, variables of left hippocampal activity and percentage left IFG damage were entered as predictors, accounting for a non-significant amount of variance in RTs (*R*^2^ = .340, *F* (2, 13) = 3.345, *p* = .067). In the second stage, the interaction term between left hippocampal activity and left IFG damage was entered accounting for a significant proportion of variance (*R*^2^ = .765, *F* (1, 12) = 5.655, *p* = .012, *b* = -.508, *t* = -2.669, *p* = .02). For visualisation, predicted simple slopes were generated to display the interaction at three levels of left IFG damage, mean and one standard deviation above/below the mean (18%, 58% and 98%; Fig. 4C). Greater hippocampal activity was associated with slower responses for those with less left IFG damage, and faster responses for those with more left IFG damage.

## DISCUSSION

Whilst there is an increasing body of research on the positive efficacy of speech and language therapy for aphasia,^3,4,74–76^ there is still limited knowledge about the neurocognitive bases of both successful therapy and its variability across patients. In this reverse translation study, therefore, we examined a commonly adopted cueing-based naming treatment and used post-intervention fMRI to explore two core questions: (1) does word re-learning in post-stroke aphasia engage the Complementary Learning Systems (CLS)^27^ that support word learning in healthy older adults?; and (2) does the level of damage to the core IFG speech production area cause deviations away from this typical learning framework?

The study yielded clear answers to both these questions. The overall result for the patients mirrored that observed in a previous post-learning fMRI study of healthy participants who had been asked to learn unfamiliar native vocabulary^28^. Specifically, following a three-week combined cueing-based and speeded naming therapy, the patients’ naming of the relearned items engaged a combination of the cortical language regions associated with production of established vocabulary plus the hippocampal episodic system. In line with the CLS framework, the gradual shift in the division of labour across these systems aligned with the consolidation status of the items (reflected in the speed and accuracy of naming). Thus, increased hippocampal activity during the naming of treated words was associated with worse performance (i.e., slower RTs and lower accuracy) whilst, in contrast, increased activity in a critical language production area, the left IFG, was associated with quicker RTs. Taken together, these counterpointed activation profiles are consistent with the CLS proposal that new learning, or in this case relearning, is initially dependent on the MTL episodic system and then gradually shifts towards long-term consolidation within the cortical systems. The results also imply that (a) the process of correcting or cleaning-up the partially-damaged cortical representations of impaired premorbid vocabulary requires a re-engagement of the MTL episodic system, and (b) long-term vocabulary reconsolidation ultimately involves re-engagement of the same cortical language production systems as those for preserved vocabulary.

In relation to the second core study question, we found that there was a crucial deviation away from this typical CLS pattern of vocabulary recovery which depended on the level of damage to the left IFG. Thus, when there was minimal IFG damage, the patients exhibited the counterpointed IFG-hippocampal correlations with naming efficiency described above and found in healthy participants when learning new vocabulary. Yet in the context of IFG damage, patients’ naming of relearned items appears to remain reliant upon the hippocampal episodic system, such that better naming performance was then associated with increased hippocampal activation. There are at least two possible explanations for this altered pattern of results when there is more damage to the cortical language systems. The first is that there is insufficient cortical tissue left to support the longer-term consolidation of the relearned words and thus the hippocampal system has to retain sole “responsibility” for relearning. The second is that, with damaged cortical systems, more restoration work is required and this correction of the impoverished representations might also take much longer than before. Accordingly, the hippocampal contribution to relearning is both greater and may be required for much longer.

These results imply that it may be important to consider the level of damage to critical language regions during stratification of patients to therapy approach. Patients with more damage to critical language regions who are likely to be, therefore, more hippocampal dependent, may show worse poorer retention over time. Indeed, the previous investigation of new vocabulary learning in healthy participants found that higher levels of hippocampal engagement after learning was associated with poorer retention at six months.^28^ To test this hypothesis in patients, further research is needed with post-treatment testing to determine maintenance of re-learned items. Furthermore, if hippocampally-dependent participants either have less efficient learning or are unable to shift the division of labour to the damaged cortical language systems, a longer-term treatment and maintenance strategy may be crucial to treatment outcomes.

Going beyond the focus on the language dominant area and hippocampal areas, the whole brain results demonstrated an overlapping cluster of increased right posterior superior temporal gyrus (pSTG) activity for both the contrasts of already known > untreated and treated > untreated. Over half of the patients had lesion overlap within the left pSTG (9; Fig. 3C). The right pSTG is consistently activated in controls and patients with aphasia during speech production tasks.^77^ Indeed, there was right pSTG activation for controls performing the same task as that used in this study.^28^ Although typically associated with receptive auditory-language process rather than speech output, the posterior STG and neighbouring regions have been argued to be a critical part of the internal feedback systems that help to monitor and thus control the accuracy of speech production.^78^

Finally, we should note that we observed very small yet reliable performance gains in the already known and untreated item sets (though considerably smaller than on the treated vocabulary). There are important considerations regarding the source of these gains in this study. Patients were tested on all items a total of five times during the course of the study. Previous studies have shown that such re-assessment alone may be sufficient to induce small yet reliable performance increases.^79^ Other studies have suggested that generalisation to untreated items via cueing therapy may be specific to the minority of patients with relatively intact semantic processing and deficits only in the phonological encoding stages of speech production.^80^ Consistent with this alternative hypothesis, the patients in this study were generally mildly aphasic and, except for one patient, all had only minor semantic deficits.

## Conclusions

The results of this study demonstrated that word re-learning post-stroke engages the same CLS dual-system framework that is found for novel vocabulary learning in healthy participants, and shows a gradual shift in the division of representational labour from the hippocampal episodic system to the cortical systems required for speech production (e.g., IFG). However, this neurotypical pattern was moderated considerably by the degree of damage to key left hemispheric language regions. After critical levels of damage to the left IFG, better performance requires a continued and increasing engagement of the hippocampal system.

## Data Availability

All data produced in the present study are available upon reasonable request to the authors

## Abbreviations

AAL: Automated Anatomical Labelling
AFNI: Analysis of Functional NeuroImages
ATL: anterior temporal lobe
BDAE: Boston Diagnostic Aphasia Examination
BOLD: blood oxygen level dependent
BNT: Boston Naming Test
CLS: Complementary Learning Systems
FDR: false discovery rate
FSL: FMRIB Software Library
IFG: inferior frontal gyrus
IPL: inferior parietal lobe
LINDA: Lesion Identification with Neighborhood Data Analysis
SLT: speech and language therapy
MNI: Montreal Neurological Institute
MTL: medial temporal lobe
pSTG: posterior superior temporal gyrus
RISP: repeated increasingly-speeded presentation
RT: reaction time
ROI: region of interest.

## Funding

Medical Research Council National Productivity Investment Fund PhD studentship; European Research Council (GAP: 670428 - BRAIN2MIND_NEUROCOMP); Medical Research Council programme grant (MR/R023883/1) and intra-mural funding (MC_UU_00005/18).

## Competing interests

The authors report no competing interests.

## Supplementary material

**Supplementary Table 1.** Percentage damage to the regions of interest (ROI) per participant. All ROIs refer to an 8mm spherical region, except AAL L IFG which refers to the whole left IFG pars opercularis and pars triangularis.

| ROI | Patient ID |  |  |  |  |  |  |  |  |  |  |  |  |  |  |  |
| --- | --- | --- | --- | --- | --- | --- | --- | --- | --- | --- | --- | --- | --- | --- | --- | --- |
|  | 1 | 2 | 3 | 4 | 5 | 6 | 7 | 8 | 9 | 10 | 11 | 12 | 13 | 14 | 15 | 16 |
| L IFG | 84 | - | - | - | 100 | 6.8 | - | - | - | 61 | 82 | - | 22 | - | - | 55 |
| R IFG | - | - | - | - | - | - | - | - | - | - | - | - | - | - | - | - |
| L ATL | - | - | 12 | - | - | - | - | - | - | - | - | - | - | - | - | - |
| L Hipp | - | 42 | 13 | - | - | - | - | 2 | - | - | - | - | 6 | - | - | 1 |
| L IPL | 87 | - | - | - | - | - | 79 | - | - | - | - | 62 | 34 | - | - | - |
| AAL L IFG | 95 | 89 | 10 | 37 | 94 | 38 | 28 | 89 | - | 88 | 99 | 97 | 95 | - | 4 | 47 |

*Note:* “-” indicates no lesion overlap with the ROI. L, left; R, right; IFG, left inferior frontal gyrus; ATL, anterior temporal lobe; Hipp, hippocampus; IPL, inferior parietal lobe; AAL, Automated Anatomical Labelling.

## Notes

### Competing Interest Statement

The authors have declared no competing interest.

### Funding Statement

This study was funded by a Medical Research Council National Productivity Investment Fund PhD studentship; European Research Council (GAP: 670428 - BRAIN2MIND_NEUROCOMP); Medical Research Council programme grant (MR/R023883/1) and intra-mural funding (MC_UU_00005/18).

### Author Declarations

NHS research ethics committee of North West Haydock Health Research Authority gave ethical approval for this work (MREC 01/8/094)

